# Knowledge and occupational practices of beauticians and barbers in the transmission of viral hepatitis: a mixed-methods study in Volta Region of Ghana

**DOI:** 10.1101/2024.06.28.24309659

**Authors:** Silas Adjei-Gyamfi, Felix Kwame Korang, Abigail Asirifi, Clotilda Asobuno

## Abstract

**Background:** Although Ghana is endemic for hepatitis B virus (HBV) and hepatitis C virus (HCV) infections, the National Policy on Viral Hepatitis stipulates that there is unreliable data, limited knowledge, and deficiency in research on viral hepatitis, especially among high-risk workers in the eastern part of the country. This study therefore assessed the knowledge level and occupational practices of beauticians and barbers in the transmission of HBV and HCV in the Volta Region of Ghana.

**Methods:** A cross-sectional mixed methods study was conducted in Volta Ghana from April to June 2021. While an in-depth interview was used to collect data from five environmental health officers who were selected as key informants in the qualitative stage, structured questionnaires/checklists, and direct observations were employed to collect data from 340 beauticians and barbers in the quantitative stage. During the qualitative stage, the process of coding, and mind mapping via thematic analysis was carried out. Furthermore, descriptive and inferential analyses were performed using Stata version 17.0 at a 95% significance level in the quantitative stage.

**Results:** Most beauticians and barbers reported poor knowledge levels about HBV and HCV (67.0%), although the awareness of this viral hepatitis was high (88.2%). While almost one-third of the observed participants practiced safe occupational activities (31.5%), 29.0%, 49.4%, and 55.3% of them observed hand hygiene, wore protective clothes/gloves, and sterilized or disinfected tools after use respectively. Beauticians and barbers with higher (tertiary) education (AOR=11.4; 95%CI=1.44–27.5; p=0.021) and those with heavy workload per day (AOR=4.34; 95%CI=1.31– 14.4; p=0.016) were more likely to report good knowledge level about HBV and HCV. Additionally, beauticians were more likely to practice safe occupational activities as compared to barbers (AOR=14.2; 95%CI=4.11–28.8; p<0.001). The key informant interviews revealed that there was little or no licensing, monitoring, and training organized for beauticians and barbers.

**Conclusion:** Participants showed high awareness but limited knowledge about HBV and HCV. The general safety practices among the participants were poor. Our study results suggest possible viral transmission through the activities of beauticians and barbers which could be attributed to the lack of regulatory systems and training of beauticians and barbers.

## Introduction

Globally, hepatitis B and C viruses are among other hepatitis viruses (A, D, E) that are of greatest concern due to their high potential for epidemics, disease burden, and mortalities they cause (1,2). Approximately more than 350 million and 170 million individuals are infected with hepatitis B virus (HBV) and hepatitis C virus (HCV) respectively in Sahalien Africa (1). The prevalence of chronic HBV and HCV infections in Ghana is estimated at 15.6% and 5.4% respectively (2–4). Additionally, asymptomatic viral hepatitis (HBV and HCV) infection is found to be present in 6.9% and 1.8% respectively of the general adult population in Ho of Volta Ghana (5). Although HBV and HCV are on the rise, the needed attention has not been given to it as compared to other viral infections like human immunodeficiency virus (HIV) in Ghana. However, the World Health Organization (WHO) has revealed that HBV and HCV are about 50 and 10 times more infectious than HIV correspondingly (1).

Beauticians and barbers work to add prettiness to their customers through shaving and styling hair and beads. Most of these workers from developing countries focus more on the decoration and entertainment of their shops than the risks related to their occupation (6). They render services to most communities in Ghana ignoring that their workplaces can be sources of HBV and HCV infections through accidental cuts and abrasions from their occupational tools (7,8), especially when proper occupational practices and infection prevention and control (IPC) measures are not observed (9,10). A study conducted in Nigeria to evaluate preventive methods among commercial barbers found that 10% and 72% of the participants sterilized and disinfected their instruments (hair clippers) respectively while 52% used kerosene, an ineffective disinfectant for disinfection procedures. In the same study, the same brush was used to clean the hair clippers and also brush the customers’ hair (8). In Southern Ghana, Atallah and his colleagues observed that 77% of barbers disinfected used tools, 63% washed their hands, and 62% used protective equipment when attending to their customers (11). A similar study in the same country also reported that most beauticians and barbers diluted their disinfectant (70% alcohol) thereby reducing its sterilization ability. They also observed that the majority of the sterilizer cabinets were not working properly (12).

The ability to prevent HBV and HBC infections among beauticians and barbers as well as their customers is highly dependent on having adequate knowledge about these infections. Some studies discovered that there is an association between knowledge level about HBV and HCV transmission among beauticians or barbers to age, educational status, and working experience (6,13). A cross-sectional study in India and Pakistan found that very few barbers (7%) were aware of HBV and HCV infections and that the reuse of blades (razors) is a potential source of HBV and HCV transmission (14,15). In Central Ghana, more than half of barbers neither knew how HBV and HCV were transmitted nor believed they could get the infection at their workplaces (6,16). In contributing to the 2030 agenda of the World Health Organization by eliminating viral hepatitis via the reduction of new infections by 90% and deaths by 65% (1), the occupational activities of beauticians and barbers and their knowledge level must be of significant concern. Although transmission of HBV and HCV is publicly understood to happen through unprotected sexual intercourse (2), however, other practical routes of infection including the operations of beauticians and barbers could be a source of transmission. It calls for studies to assess the knowledge level and occupational practices of beauticians and barbers on viral hepatitis, to bring about desirable changes in the behaviour and activities of these cosmetologists, and ultimately reduce the risk of potential transmissions.

The District/Municipal Environmental Health Unit in Ghana is a decentralized established body under the Ministry of Local Government and Rural Development (MLGRD), whose services are delivered at the assembly level (metropolitan/municipal/district). Environmental health officers (EHOs) carry out diverse roles and measures for protecting public health, including enforcing environmental health legislation and assessing, and controlling environmental and occupational factors that can potentially affect health. Formerly known as health inspectors, EHOs are responsible for monitoring and enforcing standards of environmental and public health, including food hygiene, work safety, housing pollution control, and preventing environmental health conditions injurious to health (17). However, as asserted by the Ghanaian Daily Graphic Report in 2013, there is no effective regulatory scheme in Ghana to monitor and guide the activities of some high-risk workers like beauticians and barbers. Beauticians and barbers run their private businesses and are only interested in their daily income without considering any safety precautions (18). Furthermore, there is unreliable data, limited knowledge, and a deficiency in research with no established occupational safety regulations on viral hepatitis prevention and control, especially in the Volta region of Ghana (19). This study therefore sought to assess the knowledge level and occupational practices of beauticians and barbers in the transmission of HBV and HCV in the Volta region of Ghana.

## Methods and materials

### Study area

This study was conducted in the Volta Region of Ghana, which lies within longitudes 00 15’W and 10 15’E, and latitudes 60 15’N and 80 45’N with a surface area of 10,572 km^2^. It shares borders with the Oti Region in the north, the Gulf of Guinea in the South, the Eastern Region and Lake Volta in the west, and the Republic of Togo in the east. Volta region has a population of approximately 1,960,000 residing in 18 districts or municipalities (20). Agriculture is the primary source of income in the region while other technical services like plumbing, barbering, and electrical repairs are gradually on the rise due to increased urbanization (21).

### Study design

A cross-sectional mixed-methods approach, consisting of a triangulation design was identified to conduct the study involving barbers, beauticians, and EHOs from 29th April to 30th June 2021. The quantitative method was used to provide proven statistical data whereas the qualitative study offered the opportunity to provide meaning behind the numbers. As shown in Figure 1, the quantitative strand was used to assess the knowledge level and occupational practices of beauticians and barbers. The qualitative strand was employed to conduct key informant interviews for selected EHOs. The study was therefore carried out concurrently thus, a parallel study of both quantitative and qualitative strands within the same study. Triangulation analysis of the survey, observations, and key Informant Interviews were done to form a joint interpretation of the data (22).

**Figure 1.**
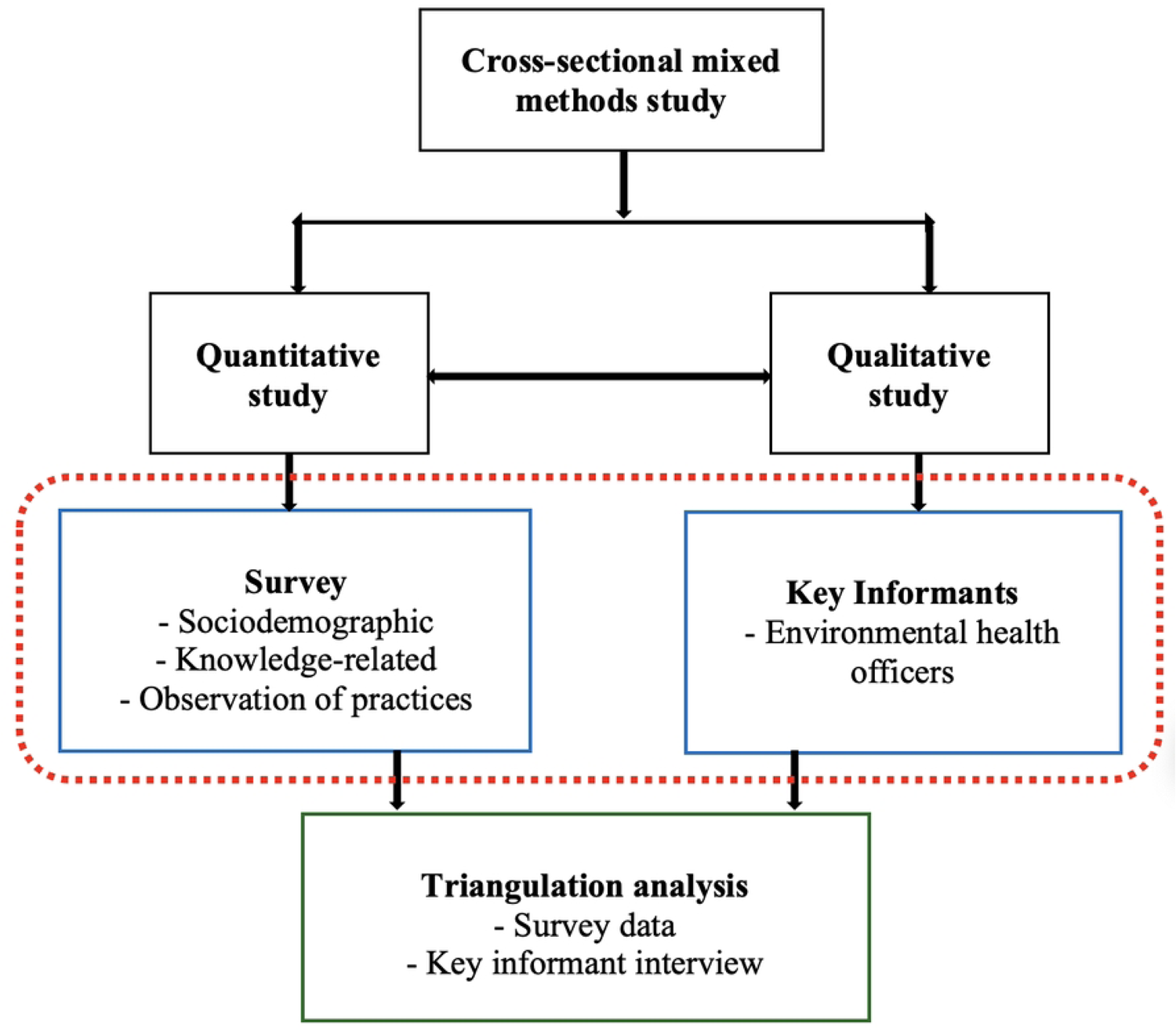
Mixed methods study design (adapted from Chandi, 2021)

### Sample size and sampling

#### Quantitative arm

Applying Cochran 1977 formula with a design effect (DEFF), the sample size (n) was primarily determined as 309 with the following indicators; prevalence (p) of HBV infection in Ghana approximated at 16% (2), margin of error (d) of 5%, standard variate 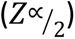 at 95% confidence level of 1.96, and DEFF of 1.5. Thus, 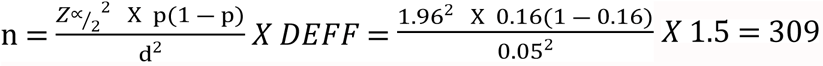. The addition of 10% attrition and non-response rate increased the estimated respondents to a final sample size of 340.

To recruit the 340 beauticians and barbers, five districts’ capital towns were selected using simple random sampling at the initial stage. The selected districts’ capital towns included Ho, Denu, Akatsi, Keta, and Sogakope which are located in Ho, Ketu South, Akatsi South, Keta, and South Tongu districts/municipals respectively. Based on the population size of each selected town, a probability proportional to size technique was employed to sample the respondents. Furthermore, due to the difficulty in locating these beauticians and barbers on account of no available member list, a snowballing sampling procedure was used to select the participants until the total sample size of 340 was attained (23,24).

#### Qualitative arm

Five senior EHOs were recruited as Key Informants using purposeful sampling. Thus, one EHO from each of the study sites (district capital). Purposeful technique was used to allow for the selection of information-rich informants who are purported to enhance understanding of beauticians’ and barbers’ practices and regulations of their practices.

### Study population

#### Quantitative arm

The study recruited 340 beauticians and barbers operating in the capital towns of selected districts of the Volta Region. This study did not include beauticians and barbers who were under 18 years old and/or were still undergoing apprenticeship.

#### Qualitative arm

This study targeted five senior and experienced EHOs from the selected district capitals of the Volta Region. However, less experienced officers like junior EHOs who have worked for one or two years were excluded from the study.

### Data collection and variables

#### Quantitative arm

A structured questionnaire containing both open-ended and closed-ended questions was used to collect data on beauticians’ and barbers’ knowledge about HBV and HCV infections and their sociodemographic characteristics. A checklist for direct observation of beauticians and barbers operating on a customer was also carried out. Informed consents were obtained from the beauticians and barbers before the observation. The practices of beauticians and barbers were observed on two occasions while attending to clients using a checklist without necessarily interacting with them. If the observation could not take place, another date was arranged with the participant. Observations usually took place between 2:00 pm and 6:00 pm on weekdays and between 1:00 pm and 6:00 pm on Saturdays and Sundays. These were the prime times the activities of the respondents could be witnessed. If multiple sessions were occurring at the same time, only one session was observed (the first session that began in the presence of the observer). Occupational practices including IPC adopted by each study participant including mode of disinfection of tools, hand washing practices, use of protective clothing, frequent use of a comb, frequency of disinfection, training on infection prevention protocols, and management of accidental cuts were observed and collected. Basic socio-demographic information including age, gender, marital status, level of education, work experience, and average number of customers per day was also collected. Participants’ knowledge level and understanding of HBV and HCV mode of transmission, signs and symptoms, and preventive measures were explored. These variables and collection tools were employed based on existing literature (8,11,16).

#### Qualitative arm

Face-to-face in-depth interviews were conducted for one senior EHO (key informant) in each study site using a semi-structured interview guide to solicit detailed information about beauticians’ and barbers’ practices as well as occupational regulations. Upon obtaining the respondents’ consent, an iPhone voice recorder was used to record the information while notes were taken during the interview. Each interview lasted for about 20 minutes.

### Statistical analysis

#### Quantitative arm

STATA version 17.0 (Stata Corporation, Texas, USA) was used to perform all analyses at a significance of p < 0.05. While knowledge, IPC, and occupational-related categorical variables were reported as frequencies and percentages, continuous variables such as age, work experience, and daily workload were presented as means and standard deviations. The outcome variables were good knowledge level and safe occupational practices whereas exposure variables included sociodemographic characteristics.

Both knowledge and practice scores were calculated for each participant. Participants received one point for each correct response to the knowledge-related questions and for each correct observed IPC and occupational practice. Incorrect responses and observations did not receive any points. By applying the median cut-off point, absolute knowledge and practice scores were estimated with their respective coefficient of Cronbach’s alpha of 0.81 and 0.79 checked to ascertain reliability. A composite knowledge score was calculated using eight items and dichotomized into good and poor knowledge levels. The practice score was also determined by using 15 correct practices and categorized into safe and unsafe occupational practices (25,26). Associations between good knowledge level or safe occupational practices and sociodemographic variables were assessed using bivariate logistic regression. Significant sociodemographic variables in the bivariate logistic regression were retained for the multivariate analyses. Multivariate (binary) logistic regression was then used to predict the factors associated with participants’ knowledge level and occupational practices.

#### Qualitative arm

The audio recordings were repeatedly listened to, translated, and transcribed by a team of experts. All note-taking was read and compared with the transcribed version to attain ideas for coding. The transcribed recordings and note-taking were then typed into Microsoft Word as the raw data for analysis after verification and review from a second opinion from the research team. Key phrases were coded and categorized by employing thematic analysis. Codes were reviewed and codes with similar meanings were grouped into working themes using mind-maps. The themes were discussed by the researchers before using them for the results writing.

#### Ethical considerations

Nagasaki University School of Tropical Medicine and Global Health Institutional Review Board (approval number: NU_TMGH_2020_130_1), and the Ghana Health Service Ethics Review Committee (approval number: GHS-ERC 036/01/21) provided ethical clearance for the study. Volta Regional Health Directorate, District/Municipal Health Directorates, and District/Municipal Assembly granted permission for this study. Written informed consent was obtained from all respondents after they were given information on the purpose of the study and what was required of them as study participants.

## Results

### Sociodemographic characteristics among respondents

Table 1 describes the social and demographic variables among the study participants. Majority of the 340 beauticians and barbers were between the ages of 20 and 29 years (61.8%) with a mean (sd) age of 26.73 (6.04) years. Although female participants (50.6%) were more than their male counterparts (49.4%), a greater proportion of the participants were single (61.8%) and had attained junior high educational status (55.9%). Less than half of these beauticians and barbers had worked for at least four years (46.8%), while less than one-third of them resided in rural communities (17.9%) and offered services to approximately 10–15 customers per day (25.0%). On the other hand, 80.0% of the EHOs were males and were more than 30 years old. More than half of the officers (60.0%) held the position of deputy district/municipal director and had working experience between 11 to 20 years.

**Table 1:** Sociodemographic characteristics of respondents.

| Variables | Category | Frequency (n) | Percentage (%) | Mean (sd) |
| --- | --- | --- | --- | --- |
| For beauticians and barbers (n = 340) |  |  |  |  |
| Age group (years) |  |  |  |  |
|  | < 20 | 32 | 9.4 | 26.73 (6.04) |
|  | 20 – 29 | 210 | 61.8 |  |
|  | 30 – 39 | 84 | 24.7 |  |
|  | 40+ | 14 | 4.1 |  |
| Gender |  |  |  |  |
|  | Male | 168 | 49.4 |  |
|  | Female | 172 | 50.6 |  |
| Marital status |  |  |  |  |
|  | Single | 210 | 61.8 |  |
|  | Married | 102 | 30.0 |  |
|  | Divorced | 10 | 2.9 |  |
|  | Cohabiting | 18 | 5.3 |  |
| Highest education level |  |  |  |  |
|  | None | 25 | 7.3 |  |
|  | Primary | 55 | 16.2 |  |
|  | Junior high | 190 | 55.9 |  |
|  | Senior high | 63 | 18.5 |  |
|  | Tertiary | 7 | 2.1 |  |
| Occupation |  |  |  |  |
|  | Beautician | 199 | 58.5 |  |
|  | Barber | 141 | 41.5 |  |
| Work experience (years) |  |  |  |  |
|  | 1 – 3 | 181 | 53.2 | 4.57 (3.86) |
|  | 4+ | 159 | 46.8 |  |
| Daily workload |  |  |  |  |
|  | < 10 persons | 241 | 70.9 | 7.89 (4.04) |
|  | 10 – 20 persons | 85 | 25.0 |  |
|  | 20+ persons | 14 | 4.1 |  |
| Place of residence |  |  |  |  |
|  | Urban | 279 | 82.1 |  |
|  | Rural | 61 | 17.9 |  |
| For environmental health officers (n = 5) |  |  |  |  |
| Age group (years) |  |  |  |  |
|  | 20 – 29 | 1 | 20.0 |  |
|  | 30 – 39 | 2 | 40.0 |  |
|  | 40+ | 2 | 40.0 |  |
| Gender |  |  |  |  |
|  | Male | 4 | 80.0 |  |
|  | Female | 1 | 20.0 |  |
| Job position |  |  |  |  |
|  | District/municipal director | 2 | 40.0 |  |
|  | Deputy district director | 3 | 60.0 |  |
| Work experience in current position (years) |  |  |  |  |
|  | 3 – 5 | 2 | 40.0 |  |
|  | 5 – 10 | 3 | 60.0 |  |
| Total work experience (years) |  |  |  |  |
|  | 3 – 5 | 1 | 20.0 |  |
|  | 5 – 10 | 1 | 20.0 |  |
|  | 11 – 20 | 3 | 60.0 |  |

### Knowledge level of hepatitis virus infections among beauticians and barbers

As illustrated in Table 2, majority of the beauticians and barbers had heard of HBV and HCV before (88.2%). Participants reported that hepatitis B and C can be transmitted through several means; 37.0% and 29.0% mentioned that it can be transmitted through unprotected sex and blood transfusion respectively. Additionally, 20% did not know how HBV and HCV can be transmitted. Almost one-third of the participants mentioned jaundice (28.0%) and muscle/joint pain (27.7%) as symptoms of HBV/HCV infections. Most of the participants did not know that HBV and HCV infections can be transmitted through sharing of their instruments (53.0%) whilst 46.7% did not believe that healthy healthy-looking persons can spread HBV or HCV. Only a few participants perceived themselves to be at risk of contracting HBV or HCV (37.1%). Hence, approximately one-third of these beauticians and barbers have a good knowledge level about HBV or HCV infections (32.9%).

**Table 2:**
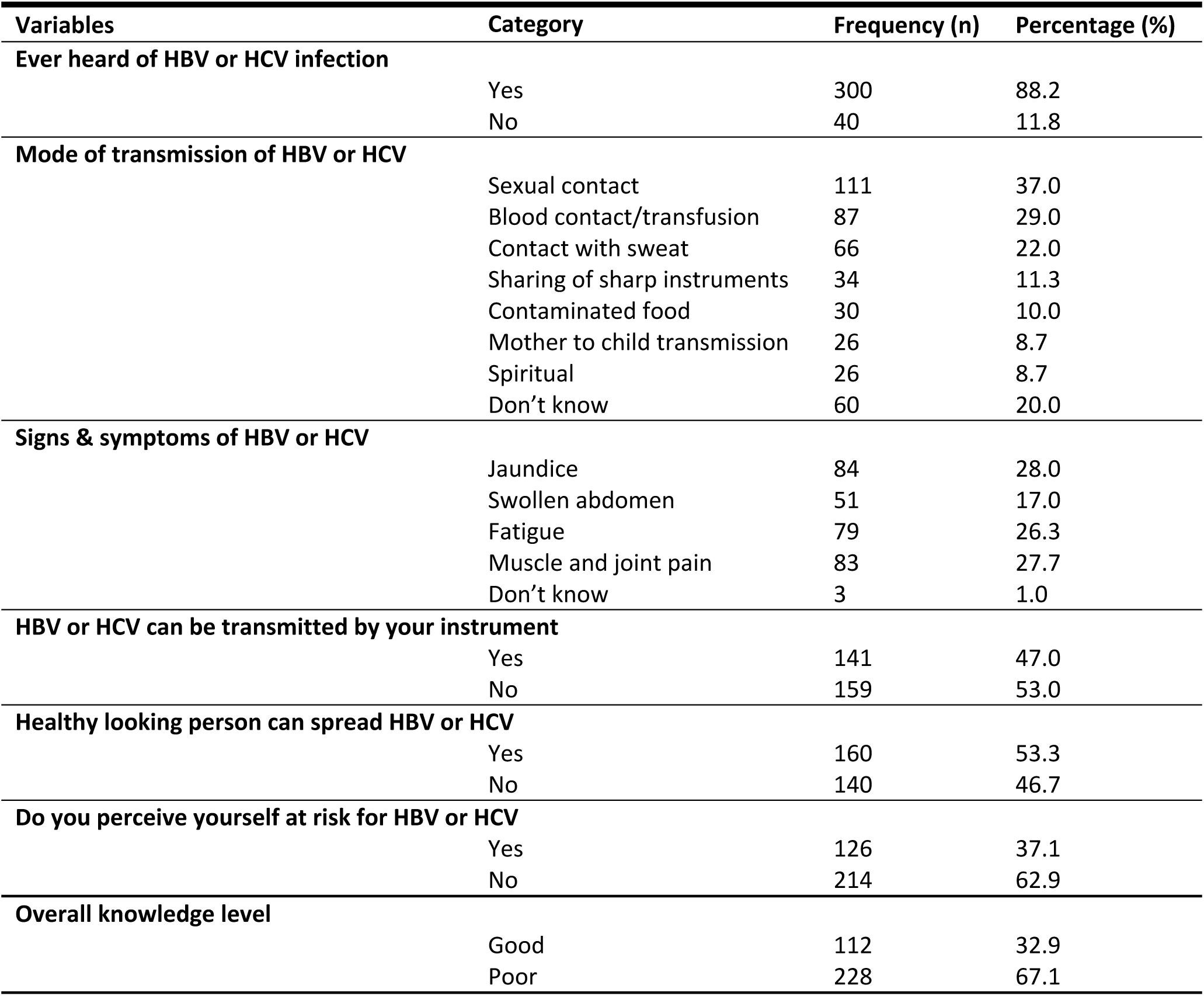
Knowledge level of hepatitis virus infection among beauticians and barbers.

### Observed occupational practices among beauticians and barbers

Table 3 shows the daily routine occupational practices including IPC among beauticians and barbers. Out of the 340 beauticians and barbers observed, none of them had a standard protocol to guide their operations whilst only 10.3% had ever received any form of training on IPC practices. A larger percentage of them used 70% alcohol-based disinfectant for disinfection procedures (73.5%) and 12.1% did not have any form of disinfectant. More than half of the participants diluted the 70% alcohol-based disinfectant before use (55.2%) and sterilized or disinfected reusable tools after using them on a customer (55.3%). Out of the 15 accidental injuries that were observed, only 40% cleaned the wound with cotton wet with alcohol.

**Table 3:**
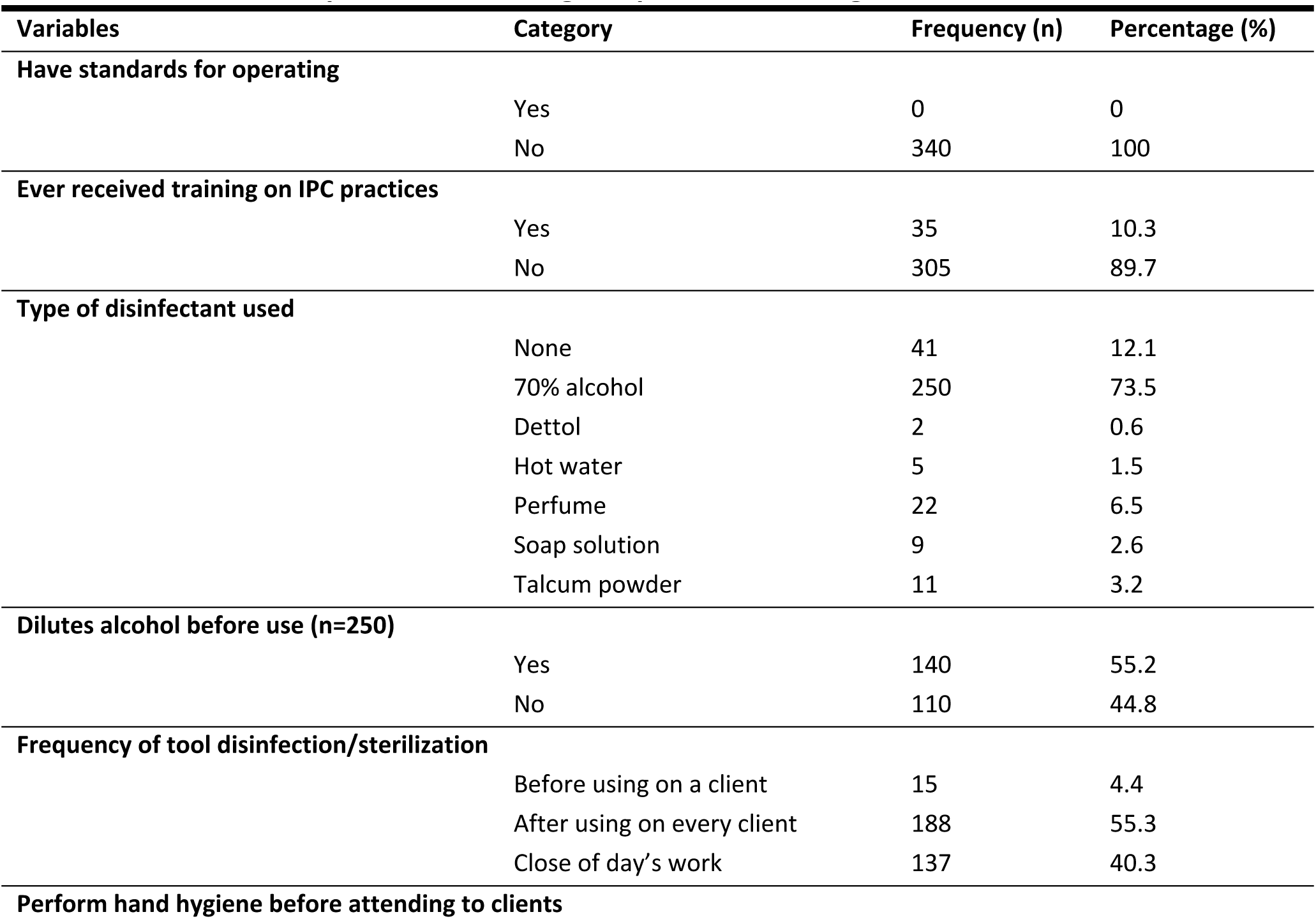
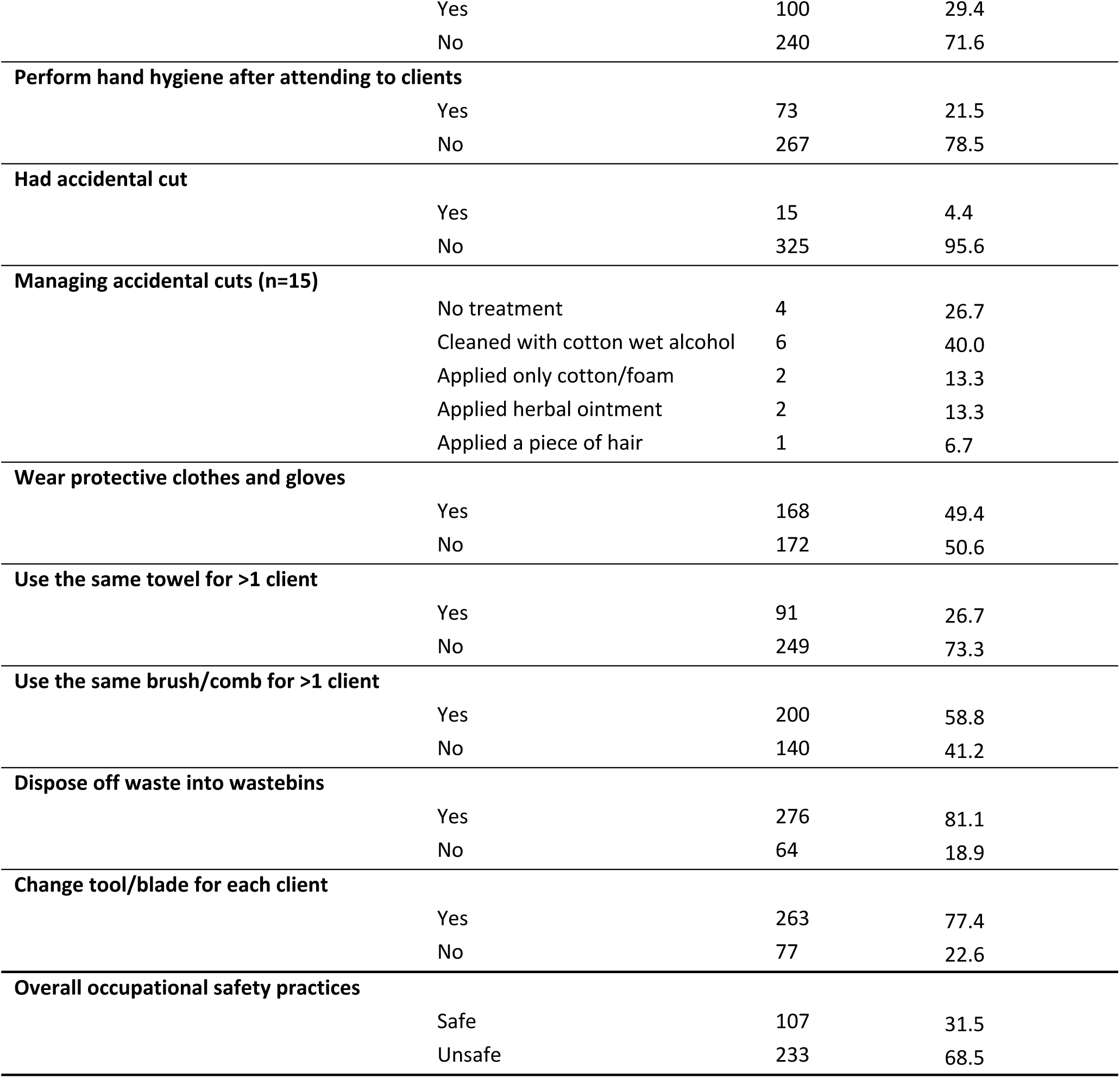
Observed occupational including IPC practices among beauticians and barbers.

Additionally, during their services with a customer, it was observed that most of the participants used a new blade or tool for every customer (77.4%), and the same hairbrush or comb for more than one customer (58.8%). While only a few used the same towel for more than one customer (26.7%) and practiced hand hygiene before attending to a customer (29.4%), majority of the participants disposed of used materials in a dustbin (81.1%). Furthermore, slightly less than half of the participants did not wear protective clothes (especially gloves) when attending to a customer (49.4%). Surprisingly, nearly one-third of the beauticians and barbers practice unsafe occupational and IPC activities (68.5%).

### Factors associated with knowledge level and occupational practices

After registering no multicollinearity among the six predictor variables (gender, marital status, educational level, occupation, work experience, daily workload) that showed significant bivariate association with good knowledge level, two of them namely educational level and daily workload remained statistically significant at the multivariate analysis level (Table 4). Moreover, as shown in Table 5, four variables including gender, educational level, occupation, and work experience were independently associated with safe occupational practices during bivariate analysis. Out of these variables, only occupation status showed a significant association in the multivariate analysis.

**Table 4:**
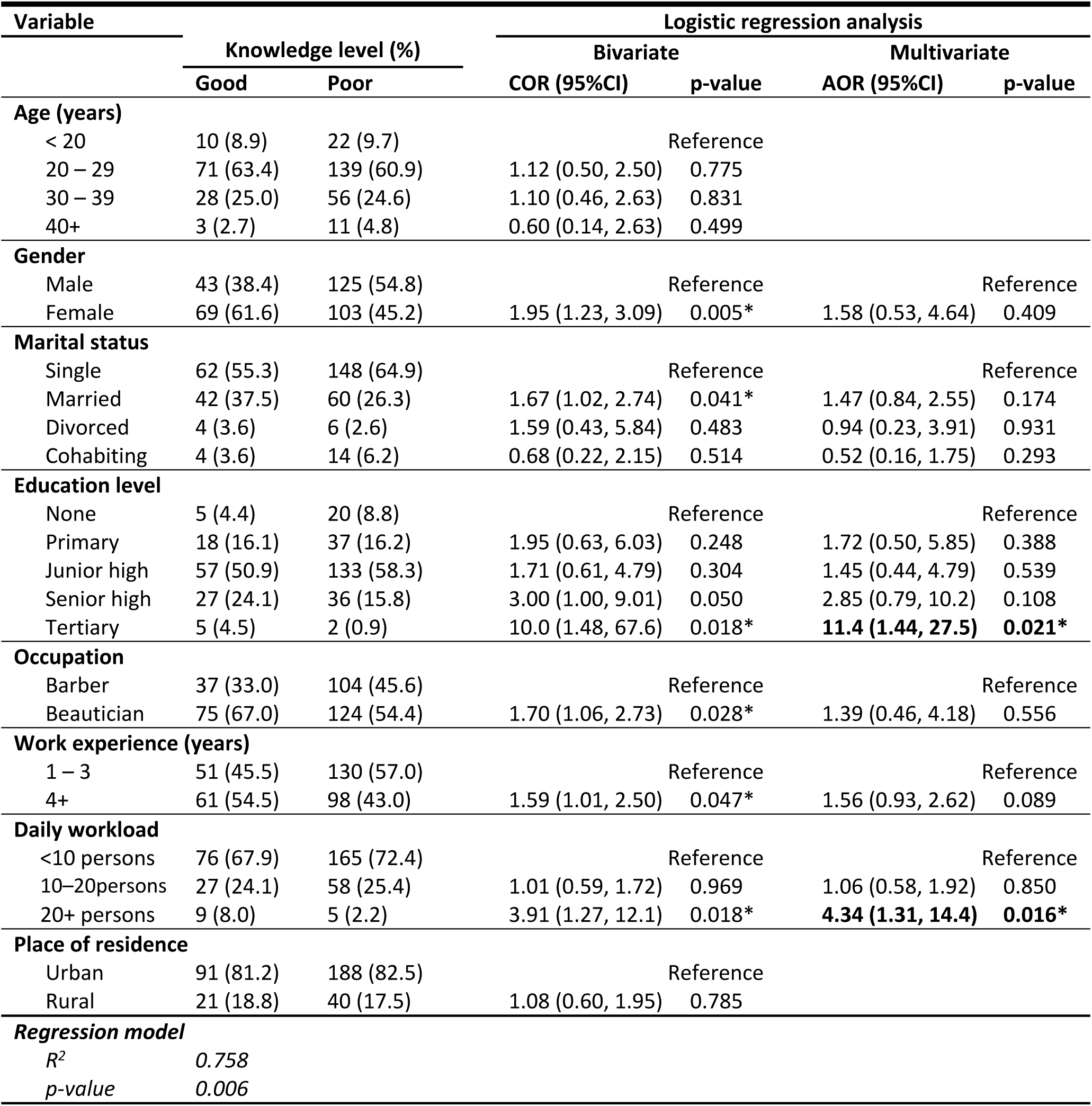
Factors associated with knowledge level.

| Variable | Logistic regression analysis |  |  |  |  |  |
| --- | --- | --- | --- | --- | --- | --- |
|  | Knowledge level (%) |  | Bivariate |  | Multivariate |  |
|  | Good | Poor | COR (95%CI) | p-value | AOR (95%CI) | p-value |
| **Age (years)** |  |  |  |  |  |  |
| < 20 | 10 (8.9) | 22 (9.7) |  | Reference |  |  |
| 20 – 29 | 71 (63.4) | 139 (60.9) | 1.12 (0.50, 2.50) | 0.775 |  |  |
| 30 – 39 | 28 (25.0) | 56 (24.6) | 1.10 (0.46, 2.63) | 0.831 |  |  |
| 40+ | 3 (2.7) | 11 (4.8) | 0.60 (0.14, 2.63) | 0.499 |  |  |
| **Gender** |  |  |  |  |  |  |
| Male | 43 (38.4) | 125 (54.8) |  | Reference |  | Reference |
| Female | 69 (61.6) | 103 (45.2) | 1.95 (1.23, 3.09) | 0.005* | 1.58 (0.53, 4.64) | 0.409 |
| **Marital status** |  |  |  |  |  |  |
| Single | 62 (55.3) | 148 (64.9) |  | Reference |  | Reference |
| Married | 42 (37.5) | 60 (26.3) | 1.67 (1.02, 2.74) | 0.041* | 1.47 (0.84, 2.55) | 0.174 |
| Divorced | 4 (3.6) | 6 (2.6) | 1.59 (0.43, 5.84) | 0.483 | 0.94 (0.23, 3.91) | 0.931 |
| Cohabiting | 4 (3.6) | 14 (6.2) | 0.68 (0.22, 2.15) | 0.514 | 0.52 (0.16, 1.75) | 0.293 |
| **Education level** |  |  |  |  |  |  |
| None | 5 (4.4) | 20 (8.8) |  | Reference |  | Reference |
| Primary | 18 (16.1) | 37 (16.2) | 1.95 (0.63, 6.03) | 0.248 | 1.72 (0.50, 5.85) | 0.388 |
| Junior high | 57 (50.9) | 133 (58.3) | 1.71 (0.61, 4.79) | 0.304 | 1.45 (0.44, 4.79) | 0.539 |
| Senior high | 27 (24.1) | 36 (15.8) | 3.00 (1.00, 9.01) | 0.050 | 2.85 (0.79, 10.2) | 0.108 |
| Tertiary | 5 (4.5) | 2 (0.9) | 10.0 (1.48, 67.6) | 0.018* | **11.4 (1.44, 27.5)** | **0.021\*** |
| **Occupation** |  |  |  |  |  |  |
| Barber | 37 (33.0) | 104 (45.6) |  | Reference |  | Reference |
| Beautician | 75 (67.0) | 124 (54.4) | 1.70 (1.06, 2.73) | 0.028* | 1.39 (0.46, 4.18) | 0.556 |
| **Work experience (years)** |  |  |  |  |  |  |
| 1 – 3 | 51 (45.5) | 130 (57.0) |  | Reference |  | Reference |
| 4+ | 61 (54.5) | 98 (43.0) | 1.59 (1.01, 2.50) | 0.047* | 1.56 (0.93, 2.62) | 0.089 |
| **Daily workload** |  |  |  |  |  |  |
| <10 persons | 76 (67.9) | 165 (72.4) |  | Reference |  | Reference |
| 10–20persons | 27 (24.1) | 58 (25.4) | 1.01 (0.59, 1.72) | 0.969 | 1.06 (0.58, 1.92) | 0.850 |
| 20+ persons | 9 (8.0) | 5 (2.2) | 3.91 (1.27, 12.1) | 0.018* | **4.34 (1.31, 14.4)** | **0.016\*** |
| **Place of residence** |  |  |  |  |  |  |
| Urban | 91 (81.2) | 188 (82.5) |  | Reference |  |  |
| Rural | 21 (18.8) | 40 (17.5) | 1.08 (0.60, 1.95) | 0.785 |  |  |
| **Regression model** |  |  |  |  |  |  |
| *R*<sup>2</sup> | 0.758 |  |  |  |  |  |
| *p*-value | 0.006 |  |  |  |  |  |

**Table 5:** Factors associated with occupational safety practices.

| Variable | Logistic regression analysis |  |  |  |  |  |
| --- | --- | --- | --- | --- | --- | --- |
|  | Occupational practices (%) |  | Bivariate |  | Multivariate |  |
|  | Safe | Unsafe | COR (95%CI) | p-value | AOR (95%CI) | p-value |
| **Age (years)** |  |  |  |  |  |  |
| < 20 | 10 (9.4) | 22 (9.4) |  | Reference |  |  |
| 20 – 29 | 77 (72.0) | 133 (57.1) | 1.27 (0.57, 2.83) | 0.553 |  |  |
| 30 – 39 | 17 (15.9) | 67 (28.8) | 0.56 (0.22, 1.39) | 0.213 |  |  |
| 40+ | 3 (2.8) | 11 (4.7) | 0.60 (0.14, 2.63) | 0.499 |  |  |
| **Gender** |  |  |  |  |  |  |
| Male | 50 (46.7) | 118 (50.6) |  | Reference |  | Reference |
| Female | 57 (53.3) | 115 (49.4) | 1.17 (1.04, 1.85) | 0.043* | 1.14 (0.44, 1.49) | 0.102 |
| **Marital status** |  |  |  |  |  |  |
| Single | 69 (64.5) | 141 (60.5) |  | Reference |  |  |
| Married | 27 (25.2) | 75 (32.2) | 0.74 (0.43, 1.24) | 0.252 |  |  |
| Divorced | 4 (3.7) | 6 (2.6) | 1.36 (0.37, 4.99) | 0.640 |  |  |
| Cohabiting | 7 (6.5) | 11 (4.7) | 1.30 (0.48, 3.50) | 0.603 |  |  |
| **Education level** |  |  |  |  |  |  |
| None | 19 (17.8) | 6 (2.6) | 8.40 (3.18, 22.2) | <0.001* | 3.04 (0.97, 9.51) | 0.057 |
| Primary | 22 (20.5) | 33 (14.2) | 1.77 (0.95, 3.31) | 0.074 | 1.35 (0.68, 2.68) | 0.393 |
| Junior high | 52 (48.6) | 138 (59.2) |  | Reference |  | Reference |
| Senior high | 12 (11.2) | 51 (21.9) | 0.62 (0.31, 1.26) | 0.191 | 0.69 (0.34, 1.42) | 0.319 |
| Tertiary | 2 (1.9) | 5 (2.1) | 1.06 (0.20, 5.64) | 0.944 | 0.94 (0.17, 5.15) | 0.940 |
| **Occupation** |  |  |  |  |  |  |
| Barber | 26 (24.3) | 115 (49.4) |  | Reference |  | Reference |
| Beautician | 81 (75.7) | 118 (50.6) | 3.04 (1.82, 5.06) | <0.001* | **14.2 (4.11, 28.8)** | **<0.001\*** |
| **Work experience (years)** |  |  |  |  |  |  |
| 1 – 3 | 62 (57.9) | 119 (51.1) |  | Reference |  | Reference |
| 4+ | 45 (42.1) | 114 (48.9) | 0.76 (0.48, 0.99) | 0.049* | 0.86 (0.52, 1.44) | 0.574 |
| **Daily workload** |  |  |  |  |  |  |
| < 10 persons | 75 (70.1) | 116 (71.2) |  | Reference |  |  |
| 10 – 15 persons | 29 (27.1) | 56 (24.1) | 1.15 (0.68, 1.93) | 0.610 |  |  |
| 20+ persons | 3 (2.8) | 11 (4.7) | 0.60 (0.16, 2.23) | 0.449 |  |  |
| **Place of residence** |  |  |  |  |  |  |
| Urban | 91 (85.0) | 188 (80.7) |  | Reference |  |  |
| Rural | 16 (15.0) | 45 (19.3) | 0.73 (0.39, 1.37) | 0.332 |  |  |
| **Regression model** |  |  |  |  |  |  |
| *R*<sup>2</sup> | 0.764 |  |  |  |  |  |
| *p*-value | < 0.001 |  |  |  |  |  |

The findings suggest that beauticians and barbers who had higher education up to the tertiary level were 11.4 times more likely to have good knowledge about HBV and HCV infections (AOR=11.4; 95%CI=1.44–27.5; p=0.021). In addition, participants who had a workload of more than 20 customers per day were 4.34 times more likely to have good knowledge about HBV and HCV infections (AOR=4.34; 95%CI=1.31–14.4; p=0.016). Beauticians were 14.2 times more likely to practice safe occupational activities as compared to barbers (AOR=14.2; 95%CI=4.11–28.8; p<0.001).

### Interview with environmental health officers

One EHO in each of the 5 study sites was interviewed to qualitatively assess the regulations of occupational safety practices of beauticians and barbers. The analysis was based on meta-narratives using thematic areas after coding.

### Core duties of environmental health officers

All the EHOs mentioned similar routine daily activities carried out by the Environmental Health Unit. However, none of the EHOs mentioned the regulation of barbers and beauticians as part of their core remit. *“…We do not specialize in one area but carry out all activities that fall within the remit of the department. First and foremost is waste disposal…, fumigation of mosquito breeding sites, routine inspection of homes to check hygiene in homes, inspection of eating and drinking areas, market hygiene… Meat inspections in abattoirs, cordoning off for dead people like COVID-19 patients, we also conduct hygiene inspections in schools, supervise zoomlion activities, and screen for food vendors. We cemetery inspectors bury the deceased, people who have died and have no owners…, we recommend building plans for approval and prosecuting environmental health violations…” EHO4*.

### Creating awareness for beauticians and barbers

All EHOs confirmed the presence and operation of barbers and beauticians in the districts/municipalities. It was also admitted that not much attention was paid to them given their practices and transmission of diseases. One respondent narrated: *“…We know that there are barbers and beauticians in the district, but we do not have much information about them because they are not under our jurisdiction. We do not inspect them like I said, we only focus on hygiene and food sanitation…” EHO3*. Another added: *“…I know about them. As part of ensuring good hygiene measures, especially for beauticians, we inspect their environment, especially in terms of how they dispose of their customers’ waste water if it has no impact on the environment. As for the barbers, since I have been in the department, we are not as strict… I can only remember us visiting the barbers once or twice to check that they had sterilized their machines…”EHO5.* One EHO mentioned that the practices of barbers and beauticians pose a very high risk to the public *“…What I can say is that the practices of some of them are very risky. They can use the same equipment for many people. Those who work in the salon are somehow better, but those who work on the street are very bad…” EHO1*.

### Licensing and regulation

In terms of licensing and regulation of beauticians and barbers in Ghana, it was found that the various districts/municipalities do not necessarily license or regulate their activities. All the EHOs indicated that there is no policy or regulatory system in place to monitor the occupational safety practices employed by beauticians and barbers. The responses of the discussants suggest that the various local assemblies are only fixated on the taxes paid by beauticians and barbers and have little or no concern for their practices. The lack of regulatory mechanisms for the activities of beauticians and barbers can lead to unhealthy practices that expose the public and themselves to blood-borne infections such as HBV and HCV. The following quotes support the findings: “… *No, we do not have any laws or policies regulating barbers. All I know is that barbers working in a kiosk have to apply for a business operating permit (BOP) at the Municipal Assembly before they start, but street barbers don’t do any registration at all… We have laws related to sanitation and food hygiene issues, but we don’t have laws for the operation of barbers…” EHO4*. Another EHO said: *“…We have no standards or laws available to us, we made bylaws to contain all social services, but they have not been gazetted what we do currently is if a beautician or a barber wants to set up a business, he/she has to come to the district assembly to take BOP.” EHO2*

### Training and continuing education

Typically, beauticians and barbers do not receive regular continuing education to be up-to-date on occupational hazards. It was evident from the responses to the discussions that no form of training was organized for them.

### Challenges among environmental health officers

One bottleneck to the inability of EHOs to monitor and enforce the safety practices of street barbers and beauticians is the lack of clear environmental regulations and laws/regulations. Participants also cited the influence of local political leaders and insufficient funds to purchase equipment and other ongoing expenses such as fuel and logistics as major obstacles to the effective monitoring of beauticians and barbers. These quotes support the views of the EHOs. *” We have problems with staff, no vehicles. We only have one motorcycle for our transportation. We fund most of our own transportation for our inspections. Regulations issued to help regulate social services have not yet been gazetted…” EHO2.* Another said: *”. Apart from the fact that there is no law to regulate barbers, we are also affected by the interference of politicians. You know, if you are a law enforcement officer and a criminal is arrested, before you take the person to prosecution, you get a call from the District Chief Executive or whoever expecting you to let the culprit go…“* EHO5

### Suggestions to initiate regulation of beauticians and barbers

Participants made some suggestions for better regulation of beauticians’ and barbers’ practices which included the provision of bylaws, policy documents, and the formation of a cosmetology association. *“…We need something like bylaws to monitor their activities, so we have something to rely on…” EHO 2. “…We should have a clearly defined policy document that regulates the activities of barbers and beauticians…” EHO3. “…As it is now, we have no idea about their total number and how to track them down because some of them are on the move. Regulating the beauticians and barbers is very difficult in the first place, I think we need to encourage them to form an association….” EHO4. “…The Assembly should provide us with the necessary resources and funding…” EHO1*.

## Discussion

Local cosmetologists commonly called beauticians and barbers serve as a major source of income for several young adults in the Volta Region of Ghana. Since there is no entry impediment to working as a beautician or barber in the country, this mixed methods study was aimed to assess the knowledge and occupational practices of beauticians and barbers in the transmission of viral hepatitis (HBV and HCV) in the Volta Region of Ghana.

### Knowledge about HBV and HCV among beauticians and barbers

Our study reported a higher level of awareness about HBV and HCV infections than the level reported among barbers in Kumasi City, Ghana (6), although most of this study’s participants have poor knowledge levels. This suggests a gradual increase in awareness of HBV and HCV infections in the Ghanaian populace. On the other hand, a significant proportion of beauticians and barbers reported wrong information about HBV and HCV transmission through contaminated food (10.0%), and spiritual means (8.7%), while 20% of them could not tell how viral hepatitis can be transmitted. Majority of the participants did not know that HBV and HCV can be transmitted by sharing instruments and did not consider themselves at risk of contracting HBV and HCV whereas less than half of them did not believe that a healthy-looking person can transmit HBV and HCV. Similarly, other studies in Ghana reported that most barbers neither knew how HBV and HCV are transmitted nor considered themselves at risk of getting these infections through their work activities (6,16). These findings show that although the majority of beauticians and barbers are aware of HBV and HCV, increased misconceptions are circulating among the Ghanaian populace.

The study further revealed that participants who had higher education up to the tertiary level were more likely to have good knowledge about HBV and HCV. Beauticians and barbers who had a workload of more than 20 customers per day were more likely to have good knowledge about HBV and HCV. These findings corroborate a study done in Pakistan, where increased workload including work experience and higher educational status were found to be associated with greater hepatitis awareness among barbers (27). Higher education may not only expose individuals to a wider range of information on infectious diseases but also prepare them to be effective, efficient, and reliable in the work environment in society (21). Literate persons are more likely to participate in outreach screening and education programs for HBV and HCV as most health promotional activities are targeted to them due to their perceived comprehension of health issues (28). Furthermore, attending to more customers per day increases workers’ performance rate and gives them more experience which consequently leads to attaining increased practical capacities and adequate knowledge about the prevention of viral hepatitis infections like HBV and HCV (29).

### Occupational practices of beauticians and barbers

With a composite occupational safety practice score of 31.5% among the study participants, beauticians were more likely to practice safe work activities as compared to barbers. This is an expected finding as most of the beauticians in our study were females and there is evidence that female beauticians effectively provide a safer work environment to their customers (30,31). The observed practices of participants showed that none of the beauticians and barbers had a protocol for infection prevention and management of accidental cuts. This study also revealed that only a few of the participants had ever received training on IPC practices. This can be attributed to a lack of oversight responsibility of the practices of beauticians and barbers by the local assemblies and relevant authorities. As Ghana continues to face a high burden of HBV and HCV infection, conscious effort needs to be devoted to IPC. In this regard, increasing the knowledge of beauticians and barbers and changing their practices is required to reduce occupational transmission. Additionally, they could be trained to serve as viral disease (HBV, and HCV) peer educators due to their unique access to the population. Although the majority of beauticians and barbers seemed to observe some form of disinfection, most of them were not performed adequately. Majority (73.5%) of the participants used 70% alcohol as a disinfectant. However, most of them (55.2%) had diluted it for economic reasons and received complaints from clients about pain sensations when undiluted alcohol-based disinfectant was applied directly to skin abrasions. This finding is consistent with a similar study in Kumasi, Ghana, which reported that most barbers diluted the 70% alcohol reducing its sterilizing ability (12). Several studies have shown that an alcohol concentration of less than 60% cannot inactivate HBV and HCV (12,32). Others used herbal ointment, soap solution, and perfume which are not known to act as disinfectants against HBV and HCV. This was an infringement of professional social responsibility to protect clients. These findings suggest that the seemingly high rate of disinfection among beauticians and barbers only gives customers and the public a false sense of safety.

Micro-traumatic injuries caused as a result of beauticians’ and barbers’ procedures on their clients can contaminate their tools, and the re-use of these tools can spread HBV and HCV (15). The study results show that 27% and 59% of participants use the same towel and comb or hair brush for each client, respectively. This could result in the effective spread of pathogens between clients through micro-abrasions.

Almost half of the participants observed used protective clothes but admitted to reusing them on other clients. Pathogens could be transferred from infected sources to used gloves and aprons, making them a potential source of transmission. The observed improper use of this protective clothing could also serve as a carrier of pathogens at the workplace (33). In terms of frequency of disinfection, a substantial number of barbers disinfected their tools at the end of the day’s work against the recommended frequency to sanitize and disinfect their tools every time between every customer (32). Further observed occupational safety practices showed that only 29.4% washed their hands before attending to a client. HBV can survive on surfaces for seven days or longer and can infect any susceptible exposed to it during that time (1). This means that the majority of the participants who did not wash their hands could pick up HBV or HCV from a customer unknowingly and possibly transmit the virus to the next.

Majority of beauticians and barbers disposed of used blades and other sharps directly into dustbins instead of recommended sharps disposal containers. This practice has continuously been posing a risk to the environment mostly to children and people who search for recycling items at the refuse dumps. It was found that the majority changed the blade or tool used by the previous customer. Due to an increase in awareness of the risk of infections through the operation of beauticians and barbers, many customers demand new razor blades or a change of tool to be used on them. However, there was no observed complaint from any customer about other unsafe practices of beauticians and barbers. Public health activities should also focus on sensitizing customers to demand safety services. The study indicated that almost one-third of the participants (31.5%) demonstrated safe practices for the prevention of HBV and HCV. This finding suggested better practices than barbers of Gondar town in Ethiopia, where 15% were reported to conduct safe practices (34). The increase in practices of participants in this study can partly be attributed to coronavirus disease 2019 (COVID-19) control measures. At the time of the data collection, it was mandatory by all workers as a social responsibility to protect themselves and the well-being of their customers amid the COVID-19 pandemic. It is also possible that participants modified their practices when they became aware that they were being observed during data collection. This suggested that risk assessment audits need to be carried out across beauticians’ and barbers’ working premises so that further information could be gathered to implement well-tailored interventions to improve their practices.

### Key informant interview

Finally, the qualitative data revealed that, unlike high-income countries where beauticians and barbers are regulated through structured training, licensing, and monitoring systems, the Government of Ghana has not given any noticeable attention to the activities of beauticians and barbers except in mobilizing taxes from them. The study found that there is no policy or regulating system to monitor the occupational safety practices employed by beauticians and barbers in the various Districts/Municipal Assemblies. This finding agrees with the report carried out by the Ghana Daily Graphic that no system monitors the operations of beauticians and barbers in Ghana (18). The participants’ poor practices can be attributed to a lack of regulatory systems and training on IPC. It is established that the provision of legislation and safety regulations is necessary for beauticians and barbers to comply with safety measures, as they provide guidance and solutions to improve health and safety at work (35).

### Limitations of the study

The participants’ perceived risk of COVID-19 among study participants and mandatory preventive measures in place at the time of data collection, such as regular hand washing and use of disinfectants, may have influenced participants’ practices. It is also possible that participants modified their practices when they became aware that they were being observed during data collection. The snowballing technique as part of the multistage sampling method used in the recruitment of study participants could lead to selection bias. Notwithstanding, the mixed methods nature of our study concurrently gives a joint interpretation of both quantitative and qualitative data which makes it unique.

## Conclusion

The study revealed a high level of awareness of HBV and HCV, however, a large proportion of participants had limited knowledge of HBV and HCV. Tertiary level of education and heavy daily workload were predictors of good knowledge level. This study found that the overall safety practices of the participants were very poor and posed a great risk for transmission of HBV and HCV through the use of ineffective disinfectants and improper handling of tools. The poor practices observed can be attributed to the lack of regulatory systems and IPC training for these beauticians and barbers. The Government of Ghana and policy-makers should institute policies and guidelines on the health and safety practices for beauticians and barbers. The Ministry of Health in collaboration with the MLGRD should organize regular obligatory practical training on IPC and workplace inspection for beauticians and barbers. The relevant regulatory agency (Department of Environmental Health) under the MLGRD should be provided with the necessary resources to monitor the activities of beauticians and barbers.

## Data Availability

The datasets collected, generated, or analyzed during this study have been attached as supplementary information.

## Acknowledgements

The authors are pleased with The Project for Human Resource Development Scholarship - Japan International Cooperation Agency (JICA); Nagasaki University School of Tropical Medicine and Global Health – Japan; and Volta Regional Health Directorate, Ghana Health Service – Ghana, for their unwavering support throughout this study. We are also grateful to all research assistants, respondents, and volunteers who greatly contributed to this study.

## Abbreviations

DEFF: Design effect
EHO: Environmental health officer
HBV: Hepatitis B virus
HCV: Hepatitis C virus
HIV: Human immunodeficiency virus
IPC: Infection prevention and control
MLGRD: Ministry of Local Government and Rural Development

## Consent For Publication

Not applicable

## Authors’ contributions

SAG, FKK, AA, and CA designed and conceptualized the study and carried out the data collection. SAG, FKK, and CA performed the data analysis and interpretation. SAG, FKK, and AA drafted the initial manuscript. All authors reviewed and approved the final manuscript.

## Authors’ acronyms

Silas Adjei-Gyamfi (SAG); Felix Kwame Korang (FKK); Abigail Asirifi (AA), and Clotilda Asobuno (CA)

## Supporting information

Figure 1. Mixed methods study design

S1 Supplementary material 1. Quantitative data

S2 Supplementary material 2. Qualitative data

